# Blood-brain barrier leakage in the penumbra is associated with infarction on follow-up imaging in acute ischemic stroke

**DOI:** 10.1101/2024.11.19.24317534

**Authors:** Felix L Nägele, Lauranne Scheldeman, Anke Wouters, Marlene Heinze, Marvin Petersen, Eckhard Schlemm, Maximilian Schell, Martin Ebinger, Matthias Endres, Jochen B Fiebach, Jens Fiehler, Ivana Galinovic, Robin Lemmens, Keith W Muir, Norbert Nighoghossian, Salvador Pedraza, Josep Puig, Claus Z Simonsen, Vincent Thijs, Götz Thomalla, Bastian Cheng

## Abstract

**Background:** Blood-brain barrier (BBB) leakage measured with dynamic susceptibility contrast magnetic resonance imaging (DSC-MRI) has been associated with hemorrhagic transformation in acute ischemic stroke (AIS). However, the influence of pre-thrombolysis BBB leakage on infarct growth has not been studied. Therefore, we aimed to characterize BBB integrity according to tissue state at admission and tissue fate on follow-up MRI.

**Methods:** This is a post-hoc analysis of the Efficacy and Safety of MRI-Based Thrombolysis in Wake-Up Stroke (WAKE-UP) trial including 165 randomized patients (mean age 66 years, 38% women, median National Institute of Health Stroke Scale score 6, 53% receiving alteplase) with available baseline DSC-MRI. Ischemic cores were segmented on diffusion weighted imaging at baseline and on fluid-attenuated inversion recovery images at follow-up (22-36h). DSC-MRI provided penumbra masks (T_max_>6s minus ischemic core) and BBB leakage (extraction fraction, EF, z-scored) maps via automated analysis. EF was averaged within the ischemic core, total penumbra, 2 penumbra subtypes (salvaged/infarcted penumbra) and normal tissue. Adjusted linear mixed effects models tested for differences between tissue types and associations of EF with clinical/imaging outcomes. Complementary voxel-wise analyses were performed.

**Results:** Linear models showed significant differences in EF between tissue types (*P*<.001). Specifically, EF was increased in the ischemic core and penumbra relative to normally-perfused tissue, while differences between total penumbra and ischemic core were statistically non-signif-icant. Penumbral tissue that later infarcted exhibited higher EF than salvaged penumbra, even after adjusting for hypoperfusion severity (*P*<.001). Voxel-wise analyses showed a significant association between EF and voxel-level infarction in the placebo group only (*P*<.001). Lesional and penumbral EF did not predict hemorrhagic transformation or functional outcomes.

**Conclusions:** Our results suggest that penumbral BBB leakage may identify tissue at increased risk of infarction. Larger, prospective studies are needed to determine the clinical relevance of BBB leakage as an imaging marker of tissue fate.

WAKE-UP http://ClinicalTrials.gov number: NCT01525290; EudraCT number: 2011-005906-32.

## INTRODUCTION

Advanced brain imaging has been crucial in identifying acute ischemic stroke (AIS) patients who are likely to benefit from intravenous thrombolysis and mechanical thrombectomy.^1^ For example, the diffusion-weighted imaging–fluid-attenuated inversion recovery (DWI-FLAIR) mismatch, serves as a ‘tissue clock’, thus facilitating the administration of intravenous thrombolysis to patients with an unknown onset of symptoms.^2^ Moreover, perfusion imaging with computed tomography (CT) and magnetic resonance imaging (MRI) leverages the pathophysiological concept of the ischemic core and penumbra^3^ and has been used successfully in the selection of AIS patients for recanalization therapies in the late time window.^4^

While recent ‘large core trials’ have challenged the need for conventional perfusion imaging in selecting patients for reperfusion therapies^5^, novel imaging techniques, such as blood-brain barrier (BBB) imaging^6^ have gained attention, as they may be useful in the study of novel treatment approaches. BBB disruption builds up within minutes to hours after stroke onset^7–9^ driven by metabolic failure, neuroinflammation, oxidative stress and increased levels of matrix metalloproteinases.^10^ Multiple studies have shown that, following the hyper acute stage (< 6 hours), BBB permeability further aggravates^9,11^, subsequently increasing the risk of hemorrhagic transformation in AIS patients.^12^ However, the relationship between pre-treatment BBB disruption and subsequent infarction or microvascular reperfusion of penumbral tissue remains unexplored. As a result, the potential application of BBB imaging to identify patients for novel treatment strategies, e.g. targeted at the neurovascular unit, has not been realized.

The present study aims to bridge this gap by characterizing BBB integrity in relation to pre-treatment tissue state and tissue fate on follow-up MRI after 22-36 hours. To achieve this, we analyzed data of patients enrolled in the WAKE-UP trial (Efficacy and Safety of MRI-Based Thrombolysis in Wake-Up Stroke) with available dynamic susceptibility contrast-enhanced (DSC)-MRI.

## METHODS

Reporting follows the STROBE (Strengthening the Reporting of Observational Studies in Epidemiology) guideline.^13^

### Study population

This post-hoc analysis includes randomized patients from the WAKE-UP trial^2^ with available baseline DSC-MRI data. The WAKE-UP trial was a multicenter, randomized, double-blind, placebo-controlled clinical trial to study the efficacy and safety of MRI-guided intravenous thrombolysis with alteplase in patients with an acute stroke of unknown time of onset. The main imaging inclusion criterion was an acute ischemic lesion visible on DWI without corresponding parenchymal hyperintensity on FLAIR, indicating that the stroke most likely had occurred within 4.5 h.

### Ethics

Patients or their legal representatives provided written informed consent according to national and local regulations. There was an exception from explicit informed consent in emergency circumstances in some countries. For each study site, the competent authorities and the corresponding ethics committee approved the trial. The trial was registered at http://ClinicalTrials.gov (NCT01525290) and EudraCT (2011-005906-32).

### Clinical data

For this analysis, we used demographic and stroke characteristics, medical history, National Institutes of Health Stroke Scale (NIHSS) and modified Rankin Scale (mRS) scores 90 days after stroke.

### Imaging data

#### Image acquisition

Brain MRI was performed on 1.5T, as well as 3T scanners according to local standards at admission and 22–36 h after randomization. Imaging sequences comprised DWI, FLAIR, susceptibility-weighted (SWI) and DSC-MRI.

#### Image processing

An overview of the image processing pipeline is shown in **Figure 1**. The analysis code comprising post-processing outside of commercial software can be accessed on GitHub: https://github.com/csi-hamburg/CSIframe/blob/main/pipelines/nice/nice.sh.

**Figure 1.**
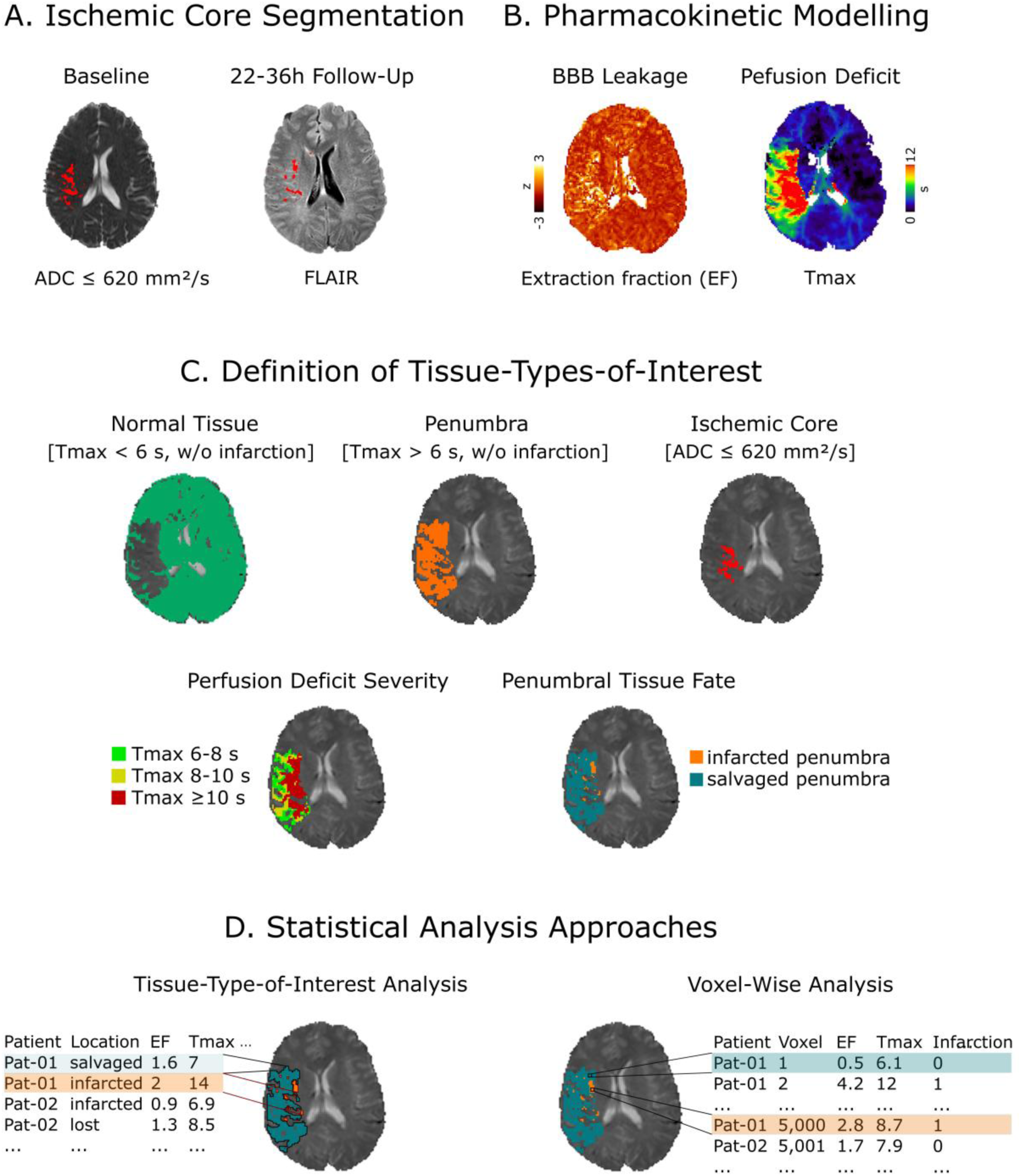
Methodological overview of the imaging pipeline. **(A)** First, ischemic cores were segmented semi-automatically on baseline apparent diffusion constant (ADC ≤620 mm²/s) and follow-up fluid-attenuated inversion recovery (FLAIR) images using the SONIA software. Then, pharmacokinetic modelling was performed in two different software suites **(B)**. 1) Nor-dicIce was used to estimate extraction fraction (EF) as a marker of blood-brain barrier (BBB) leakage, based on delay insensitive singular value deconvolution and residue function based leakage correction using an automatically defined arterial input function. 2) RAPID was used for current state of the art estimation of time to maximum of the tissue residue function (T_max_) maps and perfusion deficit masks. **(C)** After coregistration, different tissue-types-of-interest were defined using the perfusion deficit, baseline and follow-up ischemic core masks. **Panel (D)** illustrates different analytical approaches. We performed both tissue-type-of interest analyses using averaged EF/T_max_ values, and voxel-wise analyses. For simplicity only the salvaged vs. infarcted penumbra contrast is shown here.

#### Ischemic core segmentation

Baseline ischemic core lesions were segmented and quantified on apparent diffusion coefficient (ADC) maps calculated from DWI co-registered to FLAIR images using a semi-automated procedure based on an upper ADC threshold of 620 mm^2^/s (in-house Stroke Quantification Tool, SONIA, v1.0)^14,15^. The final infarct was defined as acute FLAIR hyperintensity on follow-up images (**Figure 1 Panel A**). Baseline perfusion images were linearly co-registered to FLAIR images using FLIRT (FMRIB’s Linear Image Registration Tool)^16^. The resulting inverse transformation matrices were subsequently applied to the ischemic core lesion masks.

#### Processing of DSC-MRI

We performed automated perfusion analysis utilizing the RAPID software (v4.9 and 5.0; iSchemaView, Menlo Park, CA)^17^ to derive time to maximum of the tissue residue function (T_max_) maps (**Figure 1 Panel B**), along with 4 different perfusion deficit masks: T_max_ >6s, 6-8s, 8-10s and ≥10s. In order to derive penumbra masks, baseline ischemic core lesion masks were subtracted from the T_max_>6s masks. The resulting penumbra masks were used to define infarcted penumbra and salvaged penumbra masks by multiplying or subtracting the co-registered follow-up ischemic core lesion masks, respectively (**Figure 1 Panel C**). Thus, the infarcted and salvaged penumbra regions indicate the follow-up tissue fate of the baseline penumbra.

BBB leakage was estimated as extraction fraction (EF) using NordicICE (Version 4.2.0; Nordic-NeuroLab, Bergen, Norway) via delay insensitive singular value deconvolution and residue function-based leakage correction using an automatically defined arterial input function (**Figure 1 Panel B**). EF models the ratio of permeability to perfusion, i.e. the fraction of contrast agent which reaches the extravascular space during first passage.^18,19^ The signal intensity of EF maps were z-scored to mitigate scanner differences.

Average EF and T_max_ values were extracted within different tissue-types-of-interests, i.e., the ischemic core, penumbra (total, infarcted, salvaged), perfusion deficit severities (T_max_ 6-8, 8-10 and ≥10s) and normal tissue. The latter was defined as tissue laying outside of both the ischemic core and perfusion deficit. For a complementary voxel-wise analysis, we extracted EF and T_max_ values for each voxel within the penumbra after applying a Gaussian filter (σ=0.5) to reduce noise, along with the information whether the respective voxel is part of the infarcted-penumbra (**Figure 1 Panel D**).

#### Assessment of hemorrhagic transformation

Hemorrhagic transformation was assessed 22–36h after randomization on SWI or CT, if MRI was not feasible, categorized into established radiologic subtypes according to the Heidelberg Bleeding Classification,^20,21^ and dichotomized for the present analysis in “any hemorrhagic transformation” or “no hemorrhagic transformation”. Image readings, that were blind to BBB imaging, were used from the original trial records.

### Statistical analysis

All statistical tests and plotting were performed in python (v. 3.12.1), leveraging numpy (v. 1.26.3), pandas (v. 2.14), tableone (v. 0.8.0), scipy (v. 1.11.4), pymer4 (v. 0.8.1), matplotlib (v. 3.8.0), and seaborn (v. 0.12.2). Testing was performed two-sided with *P*<.05 considered significant. Missing data were not imputed. The code is publicly available on GitHub: https://github.com/felenae/2024_bbb_wakeup.

#### Clinical and demographic data

We report descriptive statistics of the current sample with means (standard deviations), medians (inter-quartile-ranges) and counts (percentages) where applicable. We provide univariate comparisons between randomized patients receiving DSC-MRI and those who did not in the **Supplementary Material**.

#### Tissue analysis

In order to test for differences in mean EF between normal tissue, the ischemic core, and penumbra, we employed a linear mixed effects model (LME) adjusting for age, sex, baseline NIHSS and baseline ischemic core volume. Scanner and subject served as random effects. Following LMEs, Tukey post-hoc tests were performed for pairwise comparisons.

Further, an analogous LME modeled differences in EF between perfusion deficit severities (T_max_ 6-8s, T_max_ 8-10s, T_max_ ≥10s) with the same covariates and random effects, followed by Tukey post-hoc tests. This analysis was supplemented by a voxel-based LME testing the association between T_max_ and EF values within the penumbra, controlling for the same covariates, as well as random effects of T_max_ by scanner and patient.

Next, infarcted penumbra and salvaged penumbra served as tissue-types-of-interest to determine whether baseline EF differs according to tissue fate. As severe hypoperfusion has been shown to be associated with infarct progression^22^, we adjusted this model for mean T_max_ values within these areas in addition to the previously selected covariates plus treatment status (placebo vs. alteplase). The same random effects were used. Furthermore, a complementary voxel-wise analysis was performed to determine the association of penumbral EF with tissue outcome. To this end, we fitted 3 generalized LMEs with the voxel-wise binary outcome future-infarction. Voxel-wise EF, T_max_, as well as T_max_ along with EF, served as dependent variables for the first, second and third LME, respectively. Age, sex, treatment status, baseline NIHSS and baseline ischemic core-volume were included as covariates. Moreover, we included an interaction term of imaging variable(s) by treatment status to test whether EF and/or T_max_ are differentially associated with voxel-wise infarction depending on whether patients received placebo or alteplase. In case of significant interactions, post-hoc models were run for each treatment group separately. To address potential variability introduced by different scanners, and the repeated measures within each patient, random intercepts and slopes for EF and/or T_max_ by scanner and patient were included.

#### Association of EF with clinical outcomes

Our clinical outcomes of interest were dichotomized hemorrhagic transformation on follow-up MRI, excellent functional outcome (mRS 0-1) and NIHSS at 90 days. We fitted separate LMEs for EF within the baseline ischemic core and within the penumbra to predict clinical outcomes. Generalized LMEs were conducted for binary outcomes. Age, sex, baseline NIHSS, baseline ischemic core volume and treatment status served as covariates. Again, an interaction term was modelled for EF by treatment status and follow-up models were conducted for the placebo and alteplase group, separately, in case of significant interactions. To account for potential variability between different scanners, we included a random intercept for scanner and allowed the slope of EF to vary by scanner.

#### Sensitivity analysis

We fitted additional LMEs excluding subjects with missing EF data for the respective contrast of tissue analysis. For example, when investigating differences between different T_max_ thresholds, this was done only in subjects with complete data for all thresholds. Results are reported in the **Supplementary Material**.

## RESULTS

### Sample characteristics

Detailed clinical characteristics can be found in **Table 1** and **Table S1**. Of 503 patients enrolled in the clinical trial, 195 individuals received DSC-MRI. After visual quality control and image processing, 165 patients with a mean age of 65.5 years (37.6% women) had data suitable for the present analysis. Note that only 92 patients had penumbral tissue identified by RAPID; of those, 79 patients had follow-up MRI to delineate the final infarct – necessary for the calculation of infarcted and salvaged penumbra masks. The median NIHSS was 6, 21.2% of patients had a large vessel occlusion, 53.3% patients received alteplase and 21.2% exhibited any hemorrhagic transformation at fol-low-up, while symptomatic intracranial hemorrhages were rare (0.6-8.5% depending on the defining criteria). Demographics, cardiovascular risk factors/comorbidities and stroke characteristics were similar between patients receiving DSC-MRI and those who did not (*P*>.05).

**Table 1.**
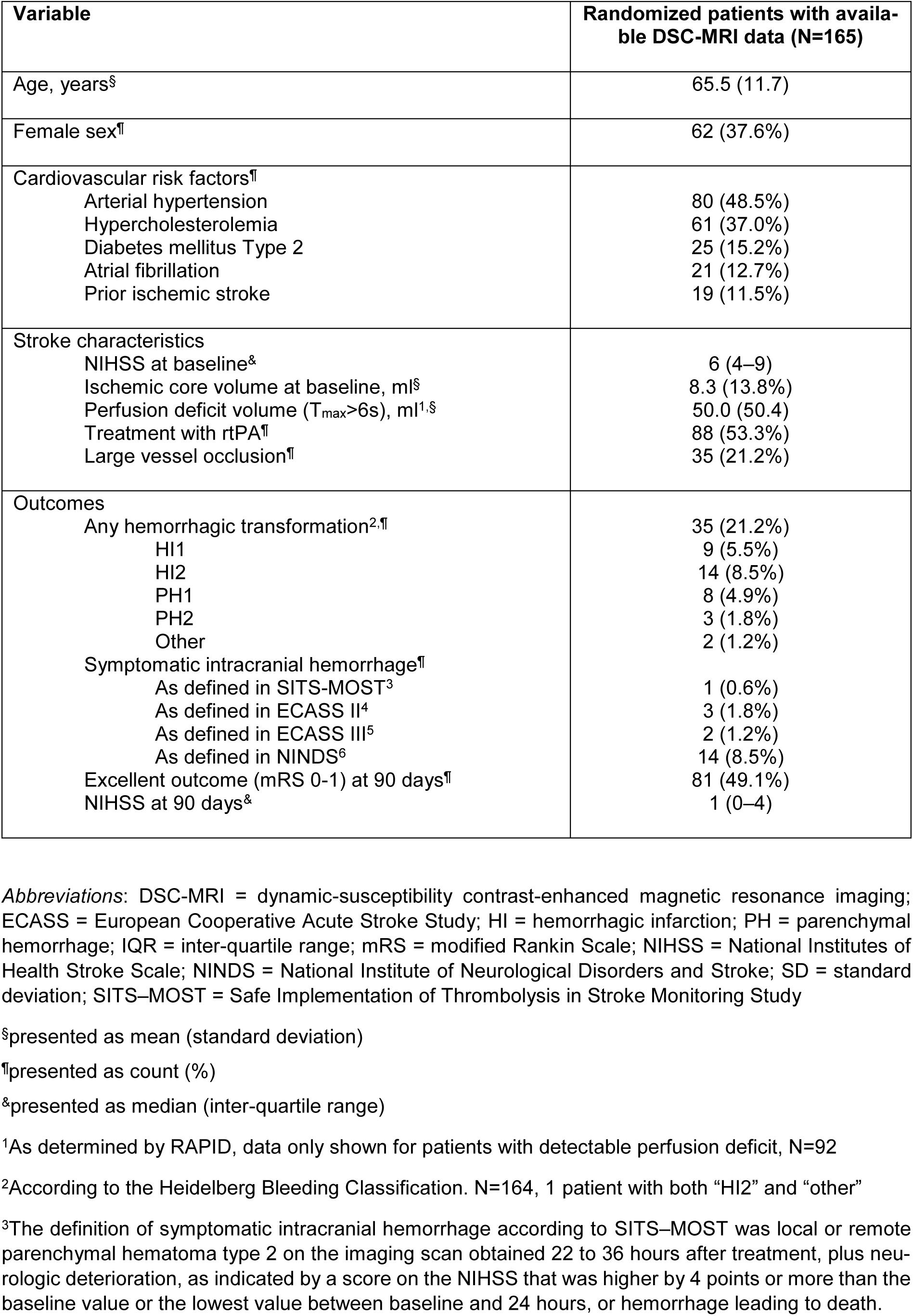
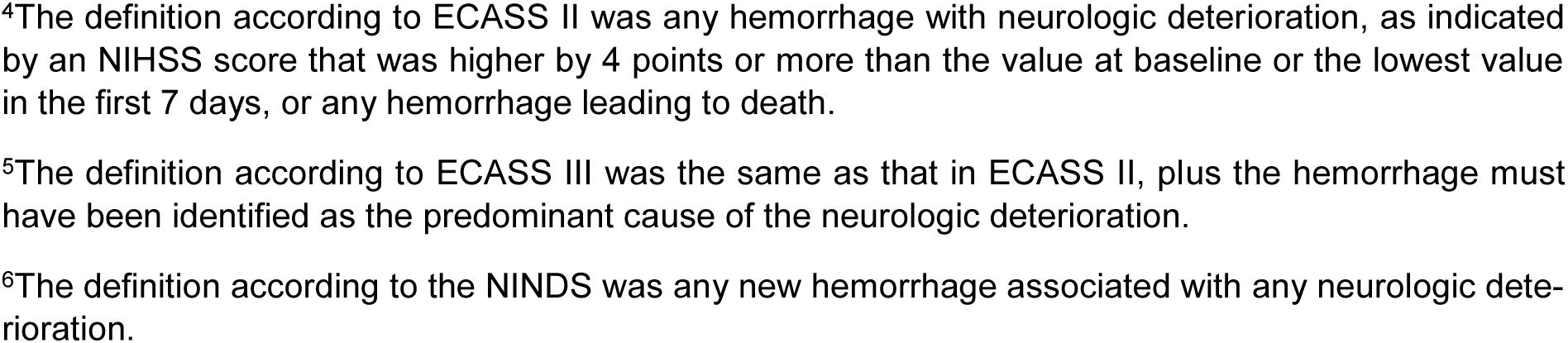
Sample characteristics.

| Variable | Randomized patients with available DSC-MRI data (N=165) |
| --- | --- |
| Age, years<sup>§</sup> | 65.5 (11.7) |
| Female sex<sup>¶</sup> | 62 (37.6%) |
| Cardiovascular risk factors<sup>¶</sup> |  |
| Arterial hypertension | 80 (48.5%) |
| Hypercholesterolemia | 61 (37.0%) |
| Diabetes mellitus Type 2 | 25 (15.2%) |
| Atrial fibrillation | 21 (12.7%) |
| Prior ischemic stroke | 19 (11.5%) |
| Stroke characteristics |  |
| NIHSS at baseline<sup>&</sup> | 6 (4–9) |
| Ischemic core volume at baseline, ml<sup>§</sup> | 8.3 (13.8%) |
| Perfusion deficit volume (T<sub>max</sub>>6s), ml<sup>1,§</sup> | 50.0 (50.4) |
| Treatment with rtPA<sup>¶</sup> | 88 (53.3%) |
| Large vessel occlusion<sup>¶</sup> | 35 (21.2%) |
| Outcomes |  |
| Any hemorrhagic transformation<sup>2,¶</sup> | 35 (21.2%) |
| HI1 | 9 (5.5%) |
| HI2 | 14 (8.5%) |
| PH1 | 8 (4.9%) |
| PH2 | 3 (1.8%) |
| Other | 2 (1.2%) |
| Symptomatic intracranial hemorrhage<sup>¶</sup> |  |
| As defined in SITS-MOST<sup>3</sup> | 1 (0.6%) |
| As defined in ECASS II<sup>4</sup> | 3 (1.8%) |
| As defined in ECASS III<sup>5</sup> | 2 (1.2%) |
| As defined in NINDS<sup>6</sup> | 14 (8.5%) |
| Excellent outcome (mRS 0-1) at 90 days<sup>¶</sup> | 81 (49.1%) |
| NIHSS at 90 days<sup>&</sup> | 1 (0–4) |

*Abbreviations:* DSC-MRI = dynamic-susceptibility contrast-enhanced magnetic resonance imaging; ECASS = European Cooperative Acute Stroke Study; HI = hemorrhagic infarction; PH = parenchymal hemorrhage; IQR = inter-quartile range; mRS = modified Rankin Scale; NIHSS = National Institutes of Health Stroke Scale; NINDS = National Institute of Neurological Disorders and Stroke; SD = standard deviation; SITS–MOST = Safe Implementation of Thrombolysis in Stroke Monitoring Study
<sup>§</sup>presented as mean (standard deviation)
<sup>¶</sup>presented as count (%)
<sup>&</sup>presented as median (inter-quartile range)
<sup>1</sup>As determined by RAPID, data only shown for patients with detectable perfusion deficit, N=92
<sup>2</sup>According to the Heidelberg Bleeding Classification. N=164, 1 patient with both “HI2” and “other”
<sup>3</sup>The definition of symptomatic intracranial hemorrhage according to SITS–MOST was local or remote parenchymal hematoma type 2 on the imaging scan obtained 22 to 36 hours after treatment, plus neurologic deterioration, as indicated by a score on the NIHSS that was higher by 4 points or more than the baseline value or the lowest value between baseline and 24 hours, or hemorrhage leading to death.
<sup>4</sup>The definition according to ECASS II was any hemorrhage with neurologic deterioration, as indicated by an NIHSS score that was higher by 4 points or more than the value at baseline or the lowest value in the first 7 days, or any hemorrhage leading to death.
<sup>5</sup>The definition according to ECASS III was the same as that in ECASS II, plus the hemorrhage must have been identified as the predominant cause of the neurologic deterioration.
<sup>6</sup>The definition according to the NINDS was any new hemorrhage associated with any neurologic deterioration.

### Tissue analysis

The LME investigating differences in EF between the non-infarcted, normally perfused tissue, acute ischemic core and penumbra showed a significant main effect of tissue-type-of-interest (EF, mean [SD]: normal = -0.15 [0.09], penumbra = 0.20 [0.38], ischemic core = 0.09 [0.41], *P*<.001; **Table 2 Part A** and **Figure 2 Panel A**). Post-hoc Tukey tests revealed that the lesion and penumbra exhibited higher EF compared to normal tissue (*P*s<.001), while there were no significant differences between the penumbra and acute ischemic core (*P*=.053).

**Figure 2.**
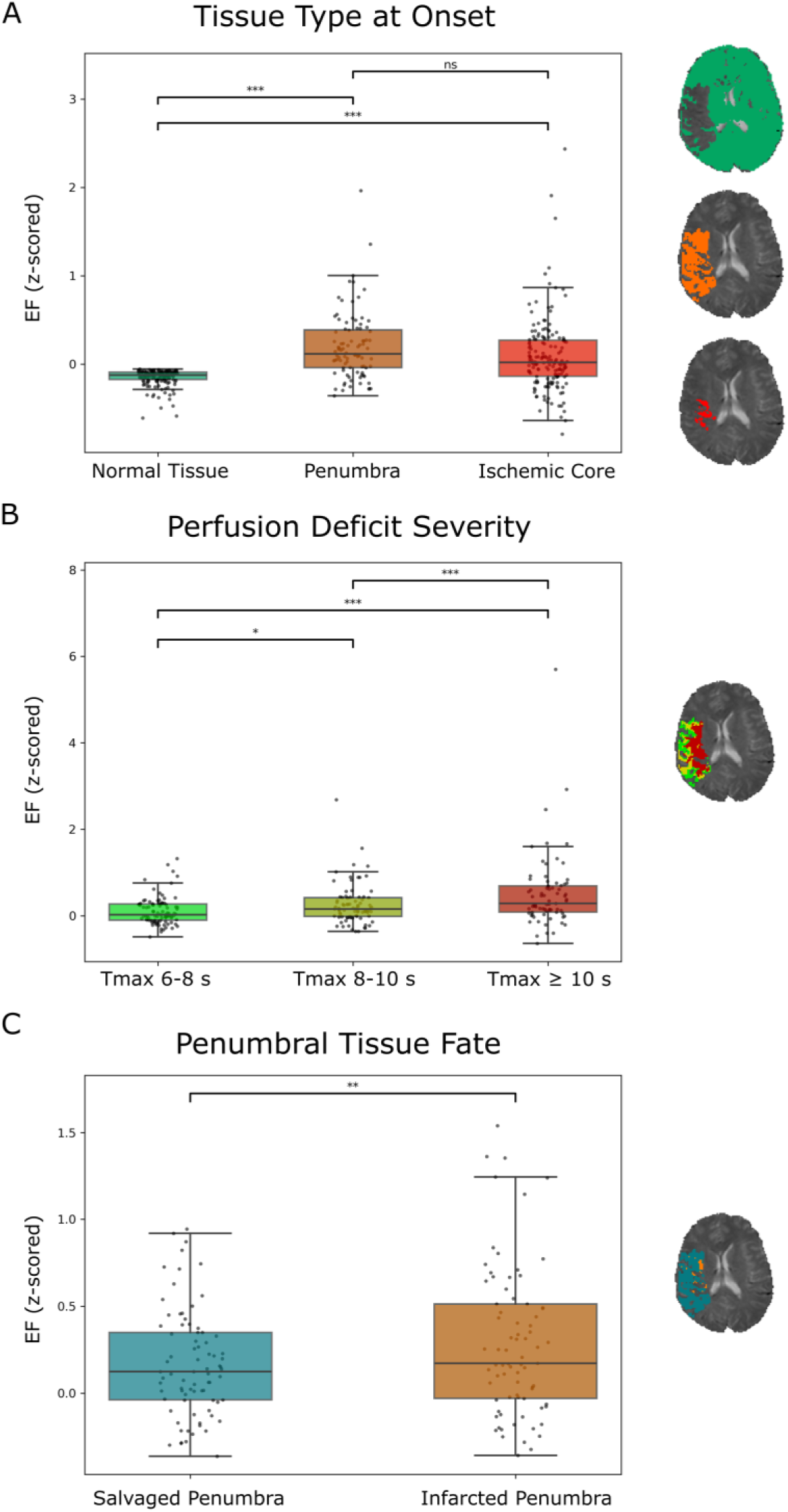
Box plots and strip plots showing the results of the linear mixed effects models and post-hoc Tukey tests. Here, differences in EF between different tissue-types of interest are illustrated with box plots and strip plots. Bars above boxplots indicate the level of significance of pair-wise post-hoc Tukey tests following adjusted linear mixed effects models. Next to each panel, schematics of the corresponding tissue-type-of-interest from Figure 1 are displayed. **Panel A** refers to the comparison of normal tissue, pe-numbra and ischemic core. **Panel B** shows differences in EF between varying T_max_ thresholds. Lastly, **Panel C** demonstrates differences in EF between the infarcted penumbra and salvaged penumbra. *Abbreviations*: DWI = diffusion weighted imaging, EF = extraction fraction, T_max_ = time to maximum of the issue residue function ^ns^non-significant; \**P*<.05; \*\**P*<.01, \*\*\**P*<.001

**Table 2.** Results of the linear mixed effects models investigating differences in EF between tissue-types-of-interest.

| Variable | Estimate [95%-CI] | T | P |
| --- | --- | --- | --- |
| A. Baseline imaging: normal tissue – ischemic core – penumbra, N=165 |  |  |  |
| EF in |  |  | **<.001<sup>\*\*\*</sup>** |
| Normal vs ischemic core<sup>#</sup> | -0.689 [-0.886; -0.491] | -8.212 | **<.001<sup>\*\*\*</sup>** |
| Normal vs penumbra<sup>#</sup> | -0.927 [-1.169; -0.685] | -9.034 | **<.001<sup>\*\*\*</sup>** |
| Ischemic core vs penumbra<sup>#</sup> | -0.239 [-0.48; 0.003] | -2.329 | .05 |
| Age | -0.037 [-0.151; 0.077] | -0.637 | .53 |
| Sex | 0.175 [-0.052; 0.401] | 1.513 | .13 |
| NIHSS | 0.047 [-0.086; 0.181] | 0.695 | .49 |
| Stroke volume | 0.037 [-0.095; 0.17] | 0.552 | .58 |
| B. Severity of hypoperfusion on baseline imaging: T<sub>max</sub> 6-8s – T<sub>max</sub> 8-10s – T<sub>max</sub> ≥10s, N=92 |  |  |  |
| EF in |  |  | **<.001<sup>\*\*\*</sup>** |
| T<sub>max</sub> 6-8s vs T<sub>max</sub> 8-10s<sup>#</sup> | -0.229 [-0.441; -0.016] | -2.539 | **.03<sup>\*</sup>** |
| T<sub>max</sub> 6-8s vs T<sub>max</sub> ≥10s<sup>#</sup> | -0.651 [-0.871; -0.430] | -6.970 | **<.001<sup>\*\*\*</sup>** |
| T<sub>max</sub> 8-10s vs T<sub>max</sub> ≥10s<sup>#</sup> | -0.422 [-0.645; -0.199] | -4.478 | **<.001<sup>\*\*\*</sup>** |
| Age | -0.114 [-0.298; 0.071] | -1.208 | .23 |
| Sex | 0.133 [-0.199; 0.466] | 0.786 | .43 |
| NIHSS | -0.004 [-.209; 0.200] | -0.042 | .97 |
| Stroke volume | -0.058 [-0.264; 0.148] | -0.550 | .58 |
| C. Tissue fate (follow-up 22-36h): salvaged penumbra – infarcted penumbra, N=79 |  |  |  |
| EF in |  |  | **<.01<sup>\*\*</sup>** |
| Salvaged vs infarcted<sup>#</sup> | -0.274 [-0.465; -0.084] | -2.860 | **<.01<sup>\*\*</sup>** |
| T<sub>max</sub> | 0.014 [-0.026; 0.053] | 0.674 | .50 |
| Age | 0.052 [-0.180; 0.283] | 0.437 | .66 |
| Sex | 0.415 [-0.002; 0.832] | 1.953 | .06 |
| NIHSS | -0.148 [-0.409; 0.114] | -1.105 | .27 |
| Stroke volume | 0.047 [-0.231; 0.325] | 0.332 | .74 |
| Treatment | -0.125 [-0.539; 0.288] | -0.595 | .55 |

*Abbreviations:* CI = confidence interval; NIHSS = National Institutes of Health Stroke Scale; T<sub>max</sub> = time to maximum of the tissue residue function
**Note:** continuous variables were z-scored before being entered into the model
\*P<.05
\*\*P<.01
\*\*\*P<.001
<sup>#</sup>Results of post-hoc Tukey tests

Testing for differences in BBB leakage within the area of perfusion deficit according to the severity of hypoperfusion, we found significantly increasing EF along with longer durations of T_max_ (6-8s = 0.11 [0.32], 8-10s = 0.26 [0.46], ≥10s = 0.52 [0.86], *P*s<.05; **Table 2 Part B** and **Figure 2 Panel B**). The voxel-wise analysis confirmed a positive linear relationship between EF and T_max_ (standardized estimate [95% CI]: 0.23 [0.16; 0.30], *P*<.001; **Table S2 Part A**).

Further, penumbral tissue with infarction on follow-up imaging showed increased mean EF compared to penumbral tissue without future infarction (infarcted penumbra = 0.28 [0.43], salvaged penumbra = 0.17 [0.30], *P*<.001; **Table 2 Part C** and **Figure 2 Panel C**).

The voxel-wise analysis revealed that 20.7% percent of all penumbral voxels showed infarction on follow-up imaging. The generalized LMEs including 74 patients, along with 246,338 voxels/observations, showed a statistically significant positive association of voxel-wise EF and T_max_ with future infarction in separate models (adjusted odds ratio [95%-confidence interval, CI], EF = 1.28 [1.15; 1.42]; T_max_ = 1.87 [1.56; 2.25], *P*<.001; **Table S2 Parts B and C;** receiver operating characteristic curves are shown in **Figure S1**). This indicates that for every one standard deviation increase in EF/T_max_, the odds of infarction increase by 28/87%. In the combined model including both EF and T_max_, increased EF was still associated with higher odds of voxel infarction (EF = 1.19 [1.07, 1.34], *P*<.01; T_max_ = 1.76 [1.45; 2.14], *P*<.001; **Table S2 Part D**). Importantly, we observed significant interactions between EF and treatment status (*P*<.01), signifying that the association of EF and voxel infarction differed between groups. Follow-up models revealed that this relationship was weaker – and statistically not significant – in patients receiving alteplase (N=39) compared to those receiving placebo (N=35) (EF [alteplase] = 1.05 [0.95; 1.15], *P*=.35; EF [placebo] = 1.35 [1.16; 1.56], *P*<.001, **Table S3 Part A**). The same pattern was observed in the combined EF plus T_max_ models stratified by group (**Table S3 Part B**). There were no significant interactions of T_max_ by treatment status.

None of the covariates demonstrated a significant effect on mean or voxel-wise EF as outcome measures (*P*>.05). Finally, male sex and higher baseline ischemic core volumes were significant predictors of voxel-wise infarction in all models (*P*<.05), while treatment, age and NIHSS were not.

### Sensitivity analysis

The sensitivity analysis supported the main findings (**Table S4**). Of note, non-significant, nominal differences in EF between the lesion and penumbra reported for the main analysis diminished when including only subjects with complete data, i.e. with penumbral tissue for this model (EF, mean [SD]: normal = -0.17 [0.09], penumbra = 0.20 [0.38], ischemic core = 0.19 [0.47], *P*=0.98).

### Association of EF with clinical outcomes

Neither lesional nor penumbral EF were significant predictors of hemorrhagic transformation or functional outcomes in (generalized) LMEs (**Table S5 and S6**). In terms of covariates, higher baseline NIHSS and ischemic core volume, as well as treatment with alteplase were associated with hemorrhagic transformation (*P*<.05). Moreover, both lower baseline NIHSS and ischemic core volume was associated with excellent functional outcome. Finally, female sex, lower baseline NIHSS and smaller baseline ischemic core volumes predicted lower 90-day NIHSS (*P*<.05). There were no significant interactions between treatment status and EF measures.

## DISCUSSION

Utilizing clinical perfusion MRI along with advanced pharmacokinetic modelling, we investigated BBB permeability changes across different pathological tissue states in patients with acute ischemic stroke. Confirming previous studies, we observed BBB disruption in the ischemic core, as defined by tissue with low ADC. In addition, we provide novel evidence of increased EF within penumbral tissue that correlates with hypoperfusion severity and subsequent tissue fate. Specifically, the data suggest that the degree of BBB disruption progresses with increasing severity of hypoperfusion and, most importantly, that elevated EF independently indicates penumbral tissue at greater risk of infarction. No statistically significant associations with clinical outcomes were found. Taken together, these findings imply that BBB imaging offers pathophysiologically meaningful information in addition to hypoperfusion severity. More research is needed to probe the clinical relevance and potential utility of BBB imaging to guide innovative treatment strategies aimed at enhancing BBB integrity in AIS patients.

### Increased extraction fraction indicates tissue state and fate

We demonstrate that the BBB is disrupted in the acute phase of AIS, as indicated by increased EF in both lesional and penumbral tissue compared to normal tissue. Multiple studies using DSC, dynamic contrast-enhanced (DCE)-MRI, and CT perfusion have documented contrast extravasation in the ischemic core at various time points._9_ Contrarily, only a few human studies have focused specifically on penumbral (i.e., potentially salvageable) tissue, yielding conflicting results. For instance, Dankbaar et al.^23^ conducted a CT perfusion study that found significantly higher BBB permeability in the ischemic core and penumbra compared to non-ischemic tissue. In contrast, a study by Horsch and colleagues^24^ reported that early non-contrast CT signs were associated with BBB leakage in the ischemic core, but not in the penumbra. Our analyses contribute to our understanding of BBB disruptions in hypoperfused tissue by showing a plausible linear relationship between higher EF and increasing T_max_ in both regional and voxel-wise analyses. Notably, this is the first human study to demonstrate that acute BBB disruption is related to future infarction of penumbral tissue, with results consistent across statistical approaches (tissue-type-of-interest/voxel-based), even when controlling for effects of hypoperfusion, measured as T_max_. However, the significant interactions between treatment status and EF in our voxel-wise models warrant consideration. Post-hoc analyses revealed that the association of EF and infarction was weaker and not statistically significant in patients treated with alteplase, suggestive of the mediating role of alteplase in mechanisms leading from vessel occlusion to infarction^25,26^.

Placing our results in the context of the current understanding of the ischemic cascade^10^, it is conceivable that the observed BBB disruption is a function of the degree of hypoperfusion caused by initial vessel occlusion. BBB leakage in turn may facilitate increased immune cell infiltration which starts around 4 to 6 hours after vessel occlusion, causing further damage to the neurovascular unit through mechanisms such as reactive oxygen species production, release of matrix metalloproteinases, and pro-inflammatory cytokines.^27^ Consequently, vasogenic edema^28^ develops, potentially hindering microcirculation in addition to other factors such as swelling of endothelial cells, vasoconstriction or pericyte contraction.^29,30^ No-reflow may further exacerbate energy and oxygen depletion, creating a vicious cycle. Intravenous thrombolysis with the recombinant tissue plasminogen activator alteplase may play a dual role in these processes. While there is ample evidence from imaging and experimental studies suggesting that alteplase is associated with increased BBB leakage^12,31–33^, its efficacy in restoring blood flow to penumbral tissue might benefit BBB integrity by averting ischemia of salvageable tissue. This direct influence on the vicious cycle of ischemia may partly explain the lesser association between pre-treatment EF and future infarction in patients treated with alteplase compared to placebo.

While our results provide an intriguing perspective, they need replication in future prospective AIS trials investigating the link between hypoperfusion, BBB leakage and tissue fate, including infarction and the no-reflow phenomenon.

### Implications for novel treatment approaches

Our results support ongoing research into neuroprotective pharmaceuticals aimed at limiting BBB damage.^34^ Of note, a recent preclinical monkey study by Debatisse et al.^35^ demonstrated the potential use of advanced BBB imaging in monitoring treatment effects, showing reductions in the volume transfer constant K_trans_ after administration of ciclosporin A in a stroke reperfusion model. A complicating factor in developing neuro-protective treatments is that different components of the ischemic cascade may be involved in both damage and repair of the neurovascular unit, depending on the timing relative to ischemia onset.^10^ This suggests that it is not only critical to think about the right target, but also about the right timing. Here, the observed BBB changes likely occurred within 4.5 hours after symptom onset^2^, suggesting that interventions targeting BBB preservation may need to be applied within a limited time window to prevent detrimental downstream effects of BBB leakage such as infarction of penumbral tissue.

### Blood-brain barrier breakdown, hemorrhagic transformation, and functional outcomes

Hemorrhagic transformation can occur in up to 30-40% of AIS patients^36^, while symptomatic intracranial hemorrhage is reported in about 5% of patients treated with rtPA^37^. Unlike previous studies^12,38^, we did not observe a significant association between increased BBB leakage and hemorrhagic transformation. A likely explanation lies in differing methodologies of BBB leakage estimation including the region-of-interest in which the measurement was conducted. Here, we calculated mean EF within each tissue-type-of-interest in order to obtain an objective measure in an automated manner without selecting a region based on qualitative interpretation. On the other hand, this could have diluted observable effects considering that only few voxels may show extreme BBB leakage associated with subsequent hemorrhage. Moreover, although we tried to mitigate heterogeneity in EF values across scanners by z-scoring image intensities and conducting mixed-effects models, the multi-center/multi-scanner design might have affected our ability to uncover between subject effects.

Besides the known risk of hemorrhagic transformation conveyed by increased BBB leakage, associations with clinical, i.e. functional, outcomes are less well established. Some studies have shown benefit in incorporating CT or MRI derived BBB leakage estimates in the prediction of NIHSS or mRS^39,40^ which is conceptually in line with our finding that higher EF was found in penumbral tissue at greater risk of infarction. Nevertheless, we did not find significant associations between average pre-treatment lesional or penumbral BBB leakage and clinical outcomes, consistent with other reports.^24,41,42^ In our study, the average ischemic core volumes of patients were small. Taken together with the high rate of excellent outcomes (49%; median NIHSS 1 after 90 days) this could have obscured any relationship between BBB leakage and functional outcomes (and/or hemorrhagic transformation) potentially observable in more severely affected patients. Again, varying methodologies in terms of BBB measurements (timing in relation to stroke-onset, imaging modality, pharmacokinetic model, region-of-interest, leakage estimation method) may in part explain discrepancies with other studies. More research is needed to find an optimal approach to elucidate associations between disturbed BBB integrity and functional outcomes in AIS patients.

### Study strengths and limitations

The strengths of our study include a novel perspective of analyzing detrimental effects of BBB in a large sample of AIS patients with longitudinal imaging data, robust statistical analyses accounting for random effects, stability of results across complementary/sensitivity analyses, and a clinically feasible imaging protocol. In terms of limitations, we have to acknowledge that, despite the relatively large sample size compared to previous studies^12,39,40^, we did not find associations between BBB leakage and clinical outcomes or hemorrhagic transformation, which may be due to our measurement approach, a sample of predominantly mild strokes with small ischemic core volumes, and/or a lack of power to which noise introduced by different scanners may have contributed. Of technical note, one has to bear in mind the limitation of modelling BBB disruptions in the ischemic core which may contain areas of extremely low cerebral blood flow (i.e., where no or negligible amounts of contrast agent arrive). Further, the reliance on commercial software – owing to the absence of open-source alternatives – limits the replicability of our findings. Lastly, the specifics of the current stroke sample (unknown symptom-onset, DWI-FLAIR mismatch, small infarcts, no thrombectomy performed) may limit the generalizability of our results for other stroke populations.

## Conclusion

We demonstrate that the degree of BBB disruption in the penumbra of the ischemic core is associated with future infarction. This extends our perspective of detrimental consequences of BBB leakage in AIS, which has been focused on the risk of hemorrhagic transformation so far. Taken together, we suggest that BBB imaging provides valuable pathological insights into AIS, and underline the need for prospective studies to investigate its clinical utility.

## Data Availability

All data produced in the present study are available upon reasonable request to the authors.

## ACKNOWLEDGMENTS

We thank the patients and their families for participating in the original trial. SOURCES OF FUNDING

The WAKE-UP Trial was supported by a grant (278276) from the European Union Seventh Framework Program.

