## Supplementary Material for "Blood-brain barrier leakage in the penumbra is associated with infarction on follow-up imaging in acute ischemic stroke"

### RESULTS

#### Sample characteristics

##### Table S1. Comparison of sample characteristics between patients with and without DSC-MRI

| Variable | Randomized patients with available DSC-MRI data (N=165) | Randomized patients without available DSC-MRI data (N=338) | *P* |
|---|---|---|---|
| Age, years[§] | 65.5 (11.7) | 65.1 (11.5) | .73 |
| Female sex[¶] | 61 (37.6%) | 116 (34.3%) | .54 |
| Cardiovascular risk factors[¶] | | | |
| Arterial hypertension | 80 (48.5%) | 186 (55.0%) | .38 |
| Hypercholesterolemia | 61 (37.0%) | 117 (34.6%) | .57 |
| Diabetes mellitus Type 2 | 25 (15.2%) | 57 (16.9%) | .60 |
| Atrial fibrillation | 21 (12.7%) | 38 (11.2%) | .80 |
| Prior ischemic stroke | 19 (11.5%) | 49 (14.5%) | .24 |
| Stroke characteristics | | | |
| NIHSS at baseline[&] | 6 (4–9) | 6 (4–10) | .97 |
| Stroke volume at baseline, ml[§] | 8.3 (13.8) | 7.7 (13.2) | .68 |
| Treatment with rtPA[¶] | 88 (53.3%) | 166 (49.1%) | .43 |
| Large vessel occlusion[¶] | 35 (21.2%) | 48 (14.2%) | .06 |
| Outcomes | | | |
| Any hemorrhagic transformation[¶] | 35 (21.2%) | 83 (24.6%) | .47 |
| Symptomatic intracranial hemorrhage[¶] | | | |
| As defined in SITS-MOST[1] | 1 (0.6%) | 5 (1.5%) | .67 |
| As defined in ECASS II[2] | 3 (1.8%) | 7 (2.1%) | .99 |
| As defined in ECASS III[3] | 2 (1.2%) | 5 (1.5%) | .99 |
| As defined in NINDS[4] | 14 (8.5%) | 18 (5.3%) | .24 |
| Excellent outcome (mRS 0-1) at 90 days[¶] | 81 (49.1%) | 152 (45%) | .44 |
| NIHSS at 90 days[&] | 1 (0–4) | 1 (0-3) | .97 |

*Abbreviations*: DSC-MRI = dynamic-susceptibility contrast-enhanced magnetic resonance imaging; ECASS = European Cooperative Acute Stroke Study; IQR = inter-quartile range; mRS = modified Rankin Scale; NIHSS = National Institutes of Health Stroke Scale; NINDS = National Institute of Neurological Disorders and Stroke; SD = standard deviation; SITS–MOST = Safe Implementation of Thrombolysis in Stroke Monitoring Study

[§]presented as mean (standard deviation)

[¶]presented as count (%)

[&]presented as median (inter-quartile range)

[1]The definition of symptomatic intracranial hemorrhage according to SITS–MOST was local or remote parenchymal hematoma type 2 on the imaging scan obtained 22 to 36 hours after treatment, plus neurologic deterioration, as indicated by a score on the NIHSS that was higher by 4 points or more than the baseline value or the lowest value between baseline and 24 hours, or hemorrhage leading to death.

[2]The definition according to ECASS II was any hemorrhage with neurologic deterioration, as indicated by an NIHSS score that was higher by 4 points or more than the value at baseline or the lowest value in the first 7 days, or any hemorrhage leading to death.

[3]The definition according to ECASS III was the same as that in ECASS II, plus the hemorrhage must have been identified as the predominant cause of the neurologic deterioration.

[4]The definition according to the NINDS was any new hemorrhage associated with any neurologic deterioration

#### Tissue analysis

**Table S2. Results of the linear mixed effects models investigating voxel-wise associations of EF, $T_{max}$ and future infarction**

| Variable | Estimate [95%-CI] | *T* | *P* |
|---|---|---|---|
| A. Voxel-wise EF ~ voxel-wise Tmax, $N_{patients}$=74; $N_{voxels}$=246,338 | | | |
| Tmax | 0.232 [0.161; 0.302] | 6.467 | **<.001*****[***]** |
| Age | 0.049 [-0.017; 0.115] | 1.461 | .15 |
| Female Sex | 0.070 [-0.061; 0.202] | 1.044 | .30 |
| NIHSS | -0.009 [-0.090; 0.072] | -0.225 | .82 |
| Stroke volume | 0.019 [-0.068; 0.105] | 0.423 | .67 |
| | **Odds ratio [95%-CI]** | *Z* | *P* |
| B. Future infarction ~ voxel-wise EF, $N_{patients}$=74; $N_{voxels}$=246,338 | | | |
| EF | 1.277 [1.148; 1.420] | 4.502 | **<.001**[***] |
| Age | 1.114 [0.822; 1.511] | 0.695 | .49 |
| Female sex | 0.518 [0.300; 0.894] | -2.364 | **.02**[*] |
| NIHSS | 1.104 [0.765; 1.593] | 0.530 | .60 |
| Stroke volume | 1.971 [1.328; 2.924] | 3.368 | **<.001**[***] |
| Treatment | 0.870 [0.489; 1.549] | -0.472 | .64 |
| EF:treatment | 0.829 [0.719; 0.955] | -2.587 | **<.01**[**] |
| C. Future infarction ~ voxel-wise Tmax, $N_{patients}$=74; $N_{voxels}$=246,338 | | | |
| $T_{max}$ | 1.870 [1.555; 2.248] | 6.652 | **<.001**[***] |
| Age | 1.168 [0.845; 1.616] | 0.940 | .35 |
| Female sex | 0.524 [0.287; 0.957] | -2.102 | **.04**[*] |
| NIHSS | 0.921 [0.621; 1.367] | -0.407 | .68 |
| Stroke volume | 2.198 [1.445; 3.343] | 3.682 | **<.001**[***] |
| Treatment | 0.938 [0.503; 1.748] | -0.203 | .84 |
| $T_{max}$:treatment | 1.151 [0.906; 1.461] | 1.152 | .25 |
| D. Future infarction ~ voxel-wise EF and Tmax, $N_{patients}$=74; $N_{voxels}$=246,338 | | | |
| EF | 1.194 [1.067; 1.335] | 3.104 | **<.01**[**] |
| $T_{max}$ | 1.760 [1.448; 2.140] | 5.681 | **<.001**[***] |
| Age | 1.172 [0.851; 1.613] | 0.971 | 0.33 |
| Female sex | 0.497 [0.275; 0.900] | -2.306 | **.02*** |
| NIHSS | 0.930 [0.625; 1.385] | -0.356 | .72 |
| Stroke volume | 2.213 [1.453; 3.371] | 3.699 | **<.001**[***] |
| Treatment | 0.929 [0.506; 1.709] | -0.236 | .81 |
| EF:treatment | 0.818 [0.710; 0.943] | -2.771 | **<.01**[**] |
| $T_{max}$:treatment | 1.190 [0.933; 1.516] | 1.403 | .16 |

*Abbreviations*: AIC = Akaike information criterion, CI = confidence interval; NIHSS = National Institutes of Health Stroke Scale, $T_{max}$ = time to maximum of the tissue residue function

**Note**: continuous variables were z-scored before being entered into the model; $^{*}P<.05$; $^{**}P<.01$; $^{***}P<.001$

**Figure S1. Receiver operating characteristic curves of voxel-wise models in Table S2 (B-D)**

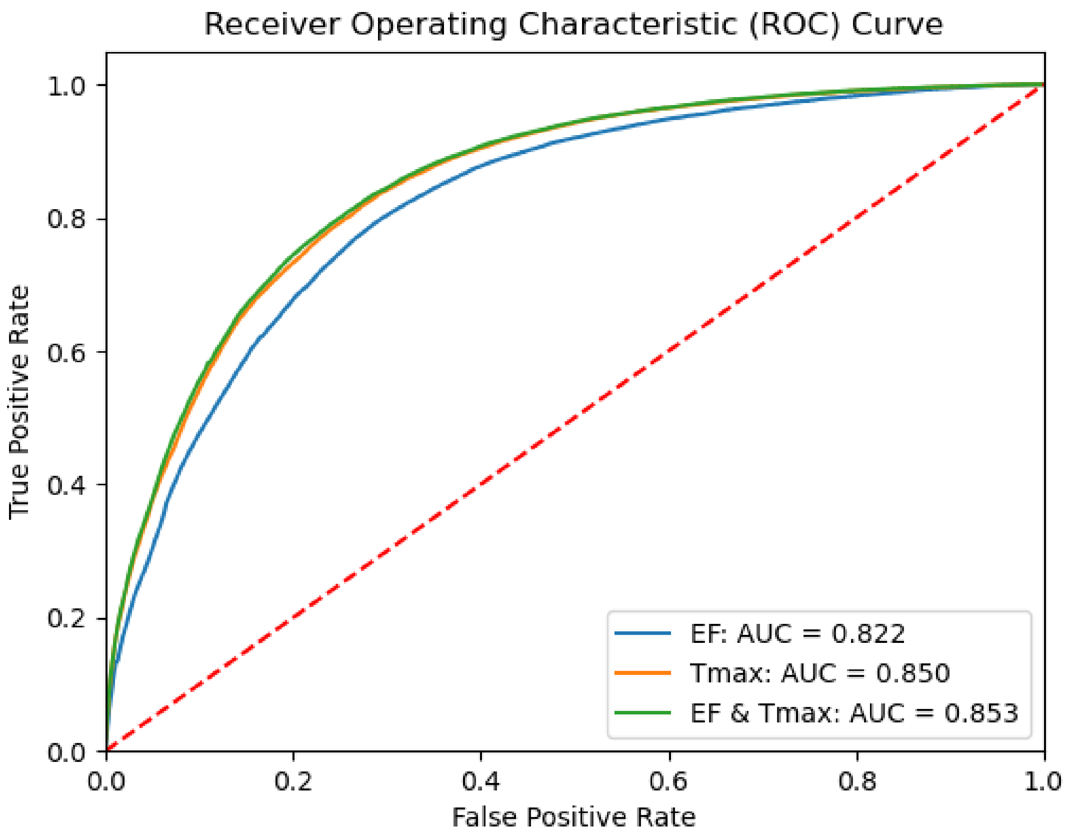

Here, receiver operating characteristic curves are shown for generalized linear mixed effects models predicting voxel-wise infarction. The curves are colored according to the imaging measures used as independent variables in the models presented in **Table S2**: blue = extraction fraction (EF, **Table S2**, **Part B**), orange = time to maximum of the tissue residue function ($T_{max}$, **Table S2**, **Part C**) and green = EF and $T_{max}$ (**Table S2**, **Part D**). The box details the mean area under the curve (AUC) which was derived by bootstrapping the sample (N=1000).

**Table S3. Results of post-hoc generalized linear mixed effects models investigating voxel-wise associations of EF, $T_{max}$ and future infarction stratified by treatment status**

| Variable | Odds ratio [95%-CI] | *Z* | *P* |
|---|---|---|---|
| A. Future infarction ~ voxel-wise EF | | | |
| **Placebo**, $N_{patients}$=35; $N_{voxels}$=108,878 | | | |
| EF | 1.345 [1.156; 1.564] | 3.839 | **<.001**$^{**}$ |
| Age | 1.478 [1.027; 2.126] | 2.104 | **.04**$^{*}$ |
| Female Sex | 0.432 [0.213; 0.874] | -2.333 | **.02**$^{*}$ |
| NIHSS | 1.223 [0.779; 1.920] | 0.874 | .38 |
| Stroke volume | 1.777 [1.189; 2.656] | 2.806 | **<.01**$^{**}$ |
| **Treatment**, $N_{patients}$=39; $N_{voxels}$=137,460 | | | |
| EF | 1.045 [0.952; 1.148] | 0.929 | .35 |
| Age | 1.015 [0.609; 1.690] | 0.056 | .96 |
| Female Sex | 0.700 [0.273; 1.795] | -0.743 | .46 |
| NIHSS | 0.935 [0.525; 1.664] | -0.228 | .82 |
| Stroke volume | 2.858 [1.297; 6.298] | 2.605 | **<.01**$^{**}$ |
| B. Future infarction ~ voxel-wise EF + $T_{max}$ | | | |
| **Placebo**, $N_{patients}$=35; $N_{voxels}$=108,878 | | | |
| EF | 1.234 [1.048; 1.454] | 2.515 | **.01**$^{*}$ |
| $T_{max}$ | 1.752 [1.388; 2.211] | 4.717 | **<.001**$^{**}$ |
| Age | 1.416 [0.975; 2.057] | 1.828 | .07 |
| Female sex | 0.351 [0.164; 0.754] | -2.687 | **<.01**$^{**}$ |
| NIHSS | 1.045 [0.650; 1.681] | 0.182 | .86 |
| Stroke volume | 1.891 [1.236; 2.892] | 2.938 | **<.01*** |
| **Treatment**, $N_{patients}$=39; $N_{voxels}$=137,460 | | | |
| EF | 0.952 [0.872; 1.041] | -1.079 | .28 |
| $T_{max}$ | 2.128 [1.761; 2.571] | 7.820 | **<.001**$^{***}$ |
| Age | 1.184 [0.703; 1.995] | 0.635 | .53 |
| Female sex | 0.631 [0.262; 1.518] | -1.029 | .30 |
| NIHSS | 0.737 [0.412; 1.319] | -1.029 | .30 |
| Stroke volume | 3.808 [1.685; 8.605] | 3.214 | **<.001**$^{***}$ |

*Abbreviations*: CI = confidence interval; NIHSS = National Institutes of Health Stroke Scale, $T_{max}$ = time to maximum of the tissue residue function

**Note**: continuous variables were z-scored before being entered into the model; $^{*}$*P*<.05; $^{***}$*P*<.001

#### Sensitivity analysis

##### Table S4. Results of the linear mixed effects models investigating differences in EF between tissue-types-of-interest without missing values

| Variable | Estimate [95%-CI] | *T* | *P* |
|---|---|---|---|
| A. Baseline imaging: normal tissue – infarct core – penumbra, N=92 | | | |
| EF in | | | **<.001*****|
| Normal *vs* infarct core# | -0.913 [-1.164; -0.662] | -8.596 | **<.001***** |
| Normal *vs* penumbra# | -0.935 [-1.186; -0.684] | -8.803 | **<.001***** |
| Infarct core *vs* penumbra# | -0.022 [-0.273; 0.229] | -0.207 | .98 |
| Age | -0.079 [-0.237; 0.078] | -0.985 | .33 |
| Sex | 0.114 [-0.180; 0.407] | 0.758 | .45 |
| NIHSS | 0.047 [-0.138; 0.232] | 0.501 | .62 |
| Stroke volume | -0.021 [-0.212; 0.171] | -0.212 | .83 |
| B. Severity of hypoperfusion: Tmax 6-8s – Tmax 8-10s – Tmax ≥10s, N=75 | | | |
| EF in | | | **<.001***** |
| Tmax 6-8s *vs* Tmax 8-10s# | -0.205 [-0.430; 0.019] | -2.163 | .08 |
| Tmax 6-8s *vs* Tmax ≥10s# | -0.608 [-0.833; -0.383] | -6.406 | **<.001***** |
| Tmax 8-10s *vs* Tmax ≥10s# | -0.403 [-0.628; -0.178] | -4.243 | **<.001***** |
| Age | -0.105 [-0.316; 0.106] | -0.976 | .33 |
| Sex | 0.163 [-0.218; 0.544] | 0.837 | .41 |
| NIHSS | -0.042 [-0.269; 0.184] | -0.364 | .72 |
| Stroke volume | -0.06 [-0.29; 0.170] | -0.512 | .61 |
| C. Tissue fate (22-36h): Salvaged penumbra – infarcted penumbra, N=77 | | | |
| EF in | | | **<.01**** |
| Salvaged *vs* infarcted# | -0.274 [-0.465; -0.083] | -2.849 | **<.01**** |
| Tmax | 0.013 [-0.027; 0.053] | 0.644 | .52 |
| Age | 0.051 [-0.181; 0.282] | 0.429 | .67 |
| Sex | 0.416 [-0.010; 0.842] | 1.912 | .06 |
| NIHSS | -0.145 [-0.409; 0.119] | -1.076 | .29 |
| Stroke volume | 0.046 [-0.233; 0.325] | 0.325 | .75 |
| Treatment | -0.126 [-0.545; 0.294] | -0.588 | .56 |

*Abbreviations*: CI = confidence interval; NIHSS = National Institutes of Health Stroke Scale; Tmax = time to maximum of the tissue residue function

**Note**: continuous variables were z-scored before being entered into the model

**$P$<.01
***$P$<.001
#Results of post-hoc Tukey tests

#### Association of EF with clinical outcomes

##### Table S5. Results of the linear mixed effects models investigating associations between lesional EF and clinical outcomes, N=165

| **Variable** | **Odds ratio/estimate [95%-CI]** | ***Z/T*** | ***P*** |
|---|---|---|---|
| | A. Excellent outcome (mRS 0-1) at 90 days¶ | | |
| EF | 1.019 [0.555; 1.872] | 0.061 | .95 |
| Age | 0.669 [0.447, 1.000] | -1.958 | .05 |
| Sex | 0.729 [0.337; 1.578] | -0.801 | .42 |
| NIHSS | 0.285 [0.153; 0.533] | -3.934 | **<.001***** |
| Stroke volume | 0.405 [0.180; 0.911] | -2.184 | **.03*** |
| Treatment | 1.524 [0.719; 3.229] | 1.100 | .27 |
| EF:treatment | 0.956 [0.447; 2.046] | -0.115 | .91 |
| | B. NIHSS at 90 days§ | | |
| EF | 0.021 [-0.139; 0.180] | 0.253 | .80 |
| Age | 0.045 [-0.059; 0.150] | 0.847 | .40 |
| Sex | -0.286 [-0.502; -0.070] | -2.597 | **.01*** |
| NIHSS | 0.543 [0.408; 0.678] | 7.875 | **<.001***** |
| Stroke volume | 0.314 [0.182; 0.447] | 4.652 | **<.001***** |
| Treatment | -0.115 [-0.320; 0.09] | -1.103 | .27 |
| EF:treatment | -0.018 [-0.224; 0.187] | -0.175 | .86 |
| | C. Hemorrhagic transformation on follow-up imaging (22-36h)¶ | | |
| EF | 1.701 [0.799; 3.619] | 1.378 | .17 |
| Age | 1.375 [0.761; 2.483] | 1.056 | .29 |
| Sex | 0.944 [0.348; 2.562] | -0.113 | .91 |
| NIHSS | 1.832 [1.029; 3.260] | 2.057 | **.04*** |
| Stroke volume | 1.701 [1.003; 2.886] | 1.970 | **.05*** |
| Treatment | 4.432 [1.561; 12.584] | 2.796 | **<.01**** |
| EF:treatment | 0.502 [0.204; 1.239] | -1.495 | .14 |

*Abbreviations*: CI = confidence interval; EF = extraction fraction; mRS = modified Rankin Scale; NIHSS = National Institutes of Health Stroke Scale

**Note**: continuous variables were z-scored before being entered into the model

**P*<.05
***P*<.01
****P*<.001
¶Estimates presented as odds ratio, along with *Z*-statistic
§Estimates presented as standardized $\beta$, along with *T*-statistic

**Table S6. Results of the linear mixed effects models investigating associations between penumbral EF and clinical outcomes, N=92**

| **Variable** | **Odds ratio/estimate [95%-CI]** | ***Z/T*** | ***P*** |
|---|---|---|---|
| A. Excellent outcome (mRS 0-1) at 90 days¶ | | | |
| EF | 1.830 [0.562; 5.957] | 1.004 | .32 |
| Age | 0.55 [0.271; 1.115] | -1.657 | .10 |
| Sex | 0.380 [0.112; 1.294] | -1.548 | .12 |
| NIHSS | 0.177 [0.058; 0.536] | -3.061 | **<.01**** |
| Stroke volume | 0.249 [0.065; 0.956] | -2.026 | **.04*** |
| Treatment | 1.348 [0.369; 4.926] | 0.452 | .65 |
| EF:treatment | 0.432 [0.112; 1.657] | -1.224 | .22 |
| B. NIHSS at 90 days§ | | | |
| EF | 0.203 [-0.059; 0.466] | 1.517 | .13 |
| Age | -0.045 [-0.181; 0.090] | -0.655 | .51 |
| Sex | -0.371 [-0.627; -0.114] | -2.832 | **<.01**** |
| NIHSS | 0.647 [0.488; 0.805] | 7.993 | **<.001***** |
| Stroke volume | 0.230 [0.071; 0.388] | 2.835 | **<.01**** |
| Treatment | -0.081 [-0.329; 0.167] | -0.641 | .52 |
| EF:treatment | -0.157 [-0.461; 0.147] | -1.014 | .32 |
| C. Hemorrhagic transformation on follow-up imaging (22-36h)¶ | | | |
| EF | 0.151 [0.009; 2.584] | -1.305 | .19 |
| Age | 1.609 [0.619; 4.181] | 0.976 | .33 |
| Sex | 0.546 [0.102; 2.926] | -0.706 | .48 |
| NIHSS | 2.667 [0.970; 7.336] | 1.900 | .06 |
| Stroke volume | 1.131 [0.481; 2.660] | 0.283 | .78 |
| Treatment | 6.252 [1.015; 38.516] | 1.976 | **<.05*** |
| EF:treatment | 1.456 [0.131; 16.209] | 0.306 | .76 |

*Abbreviations*: CI = confidence interval; EF = extraction fraction; mRS = modified Rankin Scale; NIHSS = National Institutes of Health Stroke Scale

**Note**: continuous variables were z-scored before being entered into the model

*$P$<.05
**$P$<.01
***$P$<.001
¶Estimates presented as odds ratio, along with $Z$-statistic
§Estimates presented as standardized $\beta$, along with $T$-statistic
